# A Bayesian Network Analysis of Gait Speed Change Upon Transition to Uneven Surfaces in Older Adults

**DOI:** 10.64898/2026.01.22.26344627

**Authors:** Yixiao Song, Caterina Rosano, Lana M. Chahine, Andrea L Rosso, Fabrisia Ambrosio, Nicolaas Bohnen, Seyoung Kim

## Abstract

**Background:** Gait adaptability, defined as the ability to adjust walking performance to environmental challenges, likely reflects complex interactions among the central nervous system (CNS) and other physiological systems, however, the drivers of lower gait adaptability in older adults are poorly understood.

**Methods:** We applied a Bayesian network framework to quantify multisystem interactions contributing to percent change in gait speed (%GSC) on transition from even to uneven surface in 159 older adults (63% women). Neuroimaging measures include total gray matter and white matter hyperintensities, striatal dopaminergic neurotransmission, and resting state functional connectivity. Other measures were obtained for domains important for locomotor control: health history, lifestyle, psychological well-being, cognition, and musculoskeletal and peripheral nervous systems (neurological exam). The Bayesian network estimated direct and indirect dependencies among variables, and predictive accuracy of %GSC from the Bayesian network was compared with that of multivariable linear regression using 10-fold cross-validation.

**Results:** Participants exhibited slower gait on uneven compared to even surfaces (mean %GSC = -6.32%). The Bayesian network outperformed linear regression in predicting %GSC and identified four direct paths to %GSC from: BMI, muscle strength, striato-cortical sensorimotor connectivity, and purpose in life. Indirect paths to %GSC showed interrelations among CNS and non-CNS variables, including striatal dopaminergic neurotransmission, total gray matter volume, medications, proprioception, and sex.

**Conclusions:** Gait adaptability in older adults is influenced by interactions among functional connectivity, body composition, muscle strength, and psychological well-being. Strengthening both neural and physical systems through targeted interventions may mitigate declines in gait instability and preserve mobility with aging.

## Introduction

Gait slowing is a major health concern for older adults, predicting fall risk, dementia and disabilty.^1,2^ In particular, gait slowing on surface transitions, for example from even to uneven walking surfaces, reflects adaptability to external challenges, predicts fall risks and indicates underlying neuropathology in older adults.^3–5^ Despite increasing recognition that gait speed results from interactions among central nervous system (CNS) and other physiological systems,^6–8^ research into the multisystemic nature of age-related gait slowing has been limited by traditional analytic approaches assessing one predictor at a time. These modeling approaches do not provide information on the complex interactions among predictors, posing significant challenges for understanding a multifactorial problem such as gait slowing. As an alternative, dimensionality reduction methods (e.g., principal component analysis and factor analysis) partially address the high dimensionality of predictors by identifying lower-dimensional components. However, these approaches oversimplify the complexity of gait dynamics,^9–11^ further underscoring the need for statistical methods capable of explicitly modeling the relationships among many related variables. Understanding how the CNS and other locomotor systems together influence gait speed in older age requires complex modeling approaches that have been rarely applied to studies of mobility.

Using standard linear regression modeling, we recently found that nigrostriatal dopaminergic neurotransmission is inversely related with gait slowing upon transition to uneven surface, but only in those with a high burden of peripheral systemic risk factors, such as obesity, joint pain, or reduced muscle strength.^12^ We now aim to identify the primary determinants of gait speed changes in response to uneven surfaces, while explicitly modeling the interrelationships among CNS and peripheral systemic risk factors. We apply Bayesian network modeling, a probabilistic framework well-suited for representing complex systems with dependencies among variables.^13,14^ Unlike standard regression models, which primarily focus on modeling gait speed as a function of individual predictors, a Bayesian network represents relationships among predictors as a network and combines this network with a probability model, ensuring that the probabilistic independencies align with the network structure. The probability model of the Bayesian network quantifies direct influences among the predictors and outcome, while indirect influences mediated by other variables are inferred from the model. We use the Bayesian network approach to understand the multi-factorial causes of poorer gait adaptability in older adults in the absence of clinically overt conditions.

To construct this network, we utilized comprehensive assessments of CNS and other systems important for locomotion, including body mass index (BMI), muscle strength, peripheral nervous system (PNS), joint pain, health characteristics, and lifestyle factors. The primary outcome of this study is percent decline in gait speed when transitioning from walking on an even to uneven surface, namely % gait speed change (%GSC). This measure recapitulates real-world perturbations in walking speed as may be encountered by an older adult navigating ground where the surface could change unpredictably from even to uneven, such as on a sidewalk.^5,15^

Among the CNS features, we focused on the striatal system, because of its well-known contribution to motor control and our recently shown association with gait slowing upon transitioning to uneven surfaces.^12^ The striatal system acts as a neuromodulator of mobility, via supraspinal sensorimotor networks and descending projections to the spinal cord, but also via associative and limbic (reward) networks that regulate motivation to move, executive function, and decision-making,^16–20^ as well as energy expenditure.^21^ Accordingly, we include several measures of the striatal system: striatal dopaminergic (DA) neurotransmission measured via positron emission tomography (PET); resting state functional connectivity of striatal cortical network with areas important for sensorimotor function, reward circuitry, and executive control function; and gray matter volume (GMV) of the basal ganglia.^22–30^ Markers of psychological well-being known to influence mobility were obtained via self-report using previously validated scales of motivation, purpose in life, energy and fatigability.^12,31^

We hypothesize that the Bayesian network approach will improve predictive accuracy for %GSC compared to models that evaluate individual predictors, while also revealing how these inter-related predictors influence %GSC either directly or indirectly through other variables in the network. If our results confirm the hypothesis, the Bayesian network may serve as a platform for identifying future intervention targets to preserve mobility in aging populations.

## Method

### Study Design and Participants

This neuroimaging study of mobility obtains data from participants of two ongoing epidemiological studies, the Monongahela-Youghiogheny Healthy Aging Team (MYHAT) and Study of Muscle, Mobility and Aging (SOMMA) as previously described.^32,33^

MYHAT is a community-based cohort of older adults designed to examine cognitive and functional aging. Eligible individuals were aged 65 years or older, resided in the community, and were not living in nursing homes at enrollment. Individuals were excluded if they had medical conditions or sensory impairments severe enough to prevent participation, demonstrated moderate to severe cognitive impairment based on age- and education-adjusted Mini-Mental State Examination scores below 21, or were unable to provide informed consent.

SOMMA is a prospective cohort focused on the role of muscle biology in age-related mobility decline. Eligibility criteria included age 70 years or older, residence in southwestern Pennsylvania, and the ability to walk 400 meters at a usual pace.

Exclusion criteria included use of anticoagulant medications, contraindications to magnetic resonance imaging or muscle biopsy, inability to ambulate a quarter mile or climb a flight of stairs, active malignancy, advanced chronic disease (e.g., heart failure or dialysis-dependent renal disease), Parkinson’s disease, or dementia.

For the present analyses, participants from MYHAT and SOMMA with complete data on %GSC and all variables included in the Bayesian network were retained. After excluding individuals with missing data, the final analytic sample comprised 159 participants, including 119 from SOMMA and 50 from MYHAT.

### Measure

#### Outcome

Time to complete walking of a 15-meter surface was used to compute gait speed in meters per second (m/s) under two conditions: (1) usual walking on an even surface with level flooring, free of obstacles or perturbations; (2) usual walking on an uneven surface with 1.5cm high wood prisms arranged randomly at a density of 26 pieces/m^2^ underneath carpeting.^12,34,35^

We defined our outcome %GSC as the relative change in walking speed when transitioning from an even to an uneven surface:

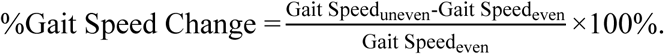

A negative value indicates a proportional slowing on uneven terrain relative to even terrain, suggesting worse adaptive capacity; values close to zero and positive values reflect preservation of gait speed, or even speeding up, when transitioning from even to uneven terrain, indicating an adaptative response.

#### Predictors

To ensure consistency and comparability across cohorts, we harmonized variables present in both datasets that are known to be related to motor function, resulting in 75 predictors (**Supplemental Table 1**).^12,36–38^ Among these, 35 variables (47%) were obtained via neuroimaging, including resting-state functional connectivity magnetic resonance imaging (MRI) measures, dopaminergic PET imaging variables, and structural MRI measures. The other 40 variables (53%) included demographics (age, sex and education), measures of musculoskeletal system (muscle strength, joint pain and fatigability), peripheral nervous systems (neurological exam), balance and parkinsonian signs (UPDRS-motor scale), as well as measures of psychological well-being (purpose in life, motivation, energy fatigability and mood), cognitive status (normal or mild cognitive impairment/dementia), lifestyle (smoking, drinking and physical activity) and general health (blood pressure, BMI, medication and pulse rate).

##### Demographic

Age, sex, education level as self-reported.

##### Musculoskeletal System

Muscle strength was assessed by maximal hand grip strength (kilograms) using a Jamar dynamometer. Participants performed two maximal-effort trials with each arm, 15 seconds apart, while seated with the arm supported and elbow flexed. The maximum value from either hand was used for analysis. Assistive walking device use was determined by asking whether the participant typically uses a walker or quad cane during ambulation. Joint pain for at least 1 month in the preceding year in the feet, toes, ankles, knees or hips was ascertained via self-report by participants.

##### Peripheral nervous system examination

A structured neurological examination conducted by trained study staff assessed downward drift or pronation of the outstretched arms. Sensory function included multiple modalities, including touch sensation, pin-prick sensation, vibration sense.

##### Motor System

Trained staff assessed motor coordination (finger-to-nose tests with eyes open and eyes closed), balance (Romberg test with eyes open and eyes closed) and parkinsonian motor features (Part III of the Movement Disorders Society-Unified Parkinson’s Disease Rating Scale, MDS-UPDRS III),^39^ which quantifies motor symptoms on an ordinal scale. Details are in **Supplemental Table 1**.

##### Psychosocial well-being factors

Motivation was measured using a reversely coded version of the self-reported 18-item Apathy Evaluation Scale (4-point Likert format), with higher scores reflecting greater motivation.^40^ Purpose in life was evaluated using the 7-item Purpose in Life subscale of Ryff’s Psychological Well-Being Scales, with higher scores indicating a stronger sense of purpose.^41^ Energy level was assessed by asking participants to rate their usual energy over the past month on a scale from 0 to 10, where 0 indicated no energy and 10 represented the highest level of energy ever experienced. Physical and mental fatigability were measured with the 10-item Pittsburgh Fatigability Scale (PFS), which quantifies anticipated fatigability following specific physical and mental activities of varying intensity on a scale from 0 (no fatigue) to 5 (extreme fatigue), scores range from 0-50 per subscale, higher score = greater fatigability.^42,43^ Mood was harmonized across cohorts using the Center for Epidemiologic Studies Depression Scale (CES-D),^44^ integrating responses from the original version in SOMMA and modified version in MYHAT, and was analyzed as a continuous variable.

##### Cognitive factor

Cognitive status was harmonized across cohorts using the Clinical Dementia Rating (CDR) in MYHAT and the Montreal Cognitive Assessment (MoCA) in SOMMA. MoCA scores were categorized as normal cognitive function, intermediate cognitive function, or low cognitive function, following harmonized percentile-based thresholds to align with CDR in MYHAT.^45^

##### Behavioral and lifestyle factors

Smoking history, alcohol intake in the prior year, and physical activity defined as number of hours of walking with at least moderate intensity per week, were reported by participants.

##### Health-related factors

BMI was defined as weight (kg) divided by height squared (m²). Seated systolic and diastolic blood pressure in mm of Hg, and average pulse (bpm) were assessed on the date of the baseline visit for each parent study; values include mean over two trials (in SOMMA) or one-time assessment (in MYHAT). Medication use reflected the total number of prescribed medications, verified by study staff from medication bottles or self-report. Vision was assessed using self-report binary items that differed slightly between cohorts: In MYHAT, participants were asked, “Do you wear your glasses or contact lenses all of the time?” whereas in SOMMA, the item asked, “Does the participant usually wear glasses or contacts for distance tasks?” Multimorbidity index captured the presence of chronic conditions, including cancer, atrial fibrillation, chronic kidney disease or renal failure, chronic obstructive pulmonary disease, myocardial infarction, heart failure, diabetes, stroke, and peripheral vascular disease.

##### Resting-state functional connectivity

Functional connectivity measures were derived from striatal subregions (executive, sensorimotor, and limbic striatum) and their cortical targets, consistent with previous work.^38^ All connectivity values represent Fisher z-transformed correlation coefficients between regional time-series extracted from preprocessed resting-state functional MRI data.

##### Brain structure

Structural MRI measures included total white matter hyperintensity (WMH) volume, normalized WMH volume, total cortical GMV, basal ganglia GMV, and brain index calculated as the ratio of total brain volume to intracranial volume. WMH quantification followed standard semi-automated segmentation pipelines. Details can be found in our previous work.^12^

##### Dopaminergic system

Dopaminergic neurotransmission was assessed using [¹¹C]dihydrotetrabenazine (DTBZ) PET to measure vesicular monoamine transporter type 2 (VMAT2) binding potential in striatal subregions: anterior putamen, pre-dorsal caudate, post-dorsal caudate, anterior ventral striatum, and posterior putamen.

Composite indices included: the volume-weighted mean of anterior putamen, pre-dorsal caudate, and post-dorsal caudate binding, and the volume-weighted mean of binding across all striatal subregions. All PET methods followed harmonized acquisition and quantification protocols described previously.^12,36–38^

### Bayesian Network: Model and Estimation

We constructed a Bayesian network that models probabilistic dependencies among the variables. A Bayesian network represents relationships among variables as a directed acyclic graph (DAG) and models a joint probability distribution over all variables that reflects the dependencies encoded in the graph. Both the graph and probability distribution were estimated from data.^46^

We set up our Bayesian network as follows. For a DAG over *N* variables *x_1_, …, x_N_*, the probability distribution associated with this graph in a Bayesian network is the product of conditional probability distributions (CPDs) for all variables:

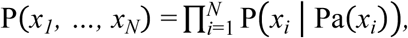

where Pa(*x_i_*) is the parents of *x_i_* defined as the set of variables with edges pointing to 𝑥_*i*_ in the graph. The CPD P(*x_i_*| Pa(*x_i_*)) for a continuous variable *x_i_* was modeled using a linear regression model, if Pa(*x_i_*) consists of continuous variables:

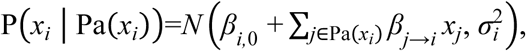

where *β_i_*,_0_, *β_j_*_→*i*_, and *σ*^2^_*i*_ are the parameters for the intercept, regression coefficient for the effect size of *x_j_* on *x_i_*, and noise variance, respectively. If any parent of the continuous variable *x_i_* is a discrete variable, we used a linear regression model for each possible value of the discrete variable. For the CPD P(*x_i_* | Pa(*x_i_*)) for discrete (binary) variables *x_i_* and Pa(*x_i_*), we used a Bernoulli distribution for each possible value of Pa(*x_i_*). In the case of a single variable in Pa(*x_i_*), we used

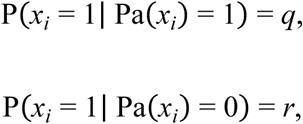

where *q* and *r* are the parameters of the Bernoulli distributions.

Both the graph structure and the parameters of the CPDs were estimated simultaneously, using the hill-climbing algorithm. The hill-climbing algorithm is an iterative optimization algorithm that, in each iteration, evaluates a set of network structure modifications and selects the one that yields the largest increase in a scoring function.^47^ A scoring function that consists of data likelihood and the Bayesian information criterion (BIC) was used to evaluate the model fit in each iteration. Since we were primarily interested in modeling how related predictors influence the outcome %GSC, in the hill-climbing algorithm we included the predictors whose Pearson correlation coefficient with the outcome is greater or equal to 0.1 and constrained the graph to have edges only among the predictors and from the predictors to the outcome.

### Overall Effects of Predictors on Outcome in Bayesian Network

While the parents of %GSC influence the outcome directly through a path with a single edge, other nodes can influence %GSC indirectly through paths that consist of multiple edges. To quantify the overall effect of each predictor on %GSC through either direct or indirect paths, we performed inference on the estimated Bayesian network and derived the overall marginal effect 𝛼*_j_* of the predictor *x_j_* on the outcome as the conditional distribution

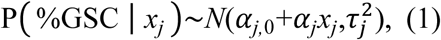

where 𝛼_*j*,0_ is the intercept, 𝛼*_j_* is the desired marginal effect of the predictor *x_j_*, and 𝜏^2^_*j*_ is the residual variance. To derive this conditional distribution, we first derived the joint probability distribution of all variables, including predictors and outcome, by topologically sorting the variables and iteratively constructing partial joint distributions over subsets of the sorted variables. From this full joint Gaussian distribution, the conditional probability distribution in Eq. (1) was obtained. For discrete variables with many downstream nodes, marginal effects were estimated separately for each value of the discrete variables.

### Path Contribution in Bayesian Network

For predictors with direct paths to %GSC, the path contribution was obtained as the regression coefficient of the corresponding predictor in the CPD of %GSC. For predictors with indirect paths to %GSC, we quantified the path contribution by decomposing the marginal effects in Eq. (1) into parts arising from individual paths to %GSC. This amounts to computing the product of the regression coefficients in the CPDs along the path:

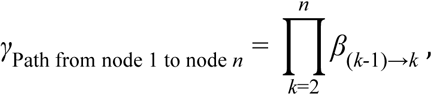

where *β*_(*k-*1)→*k*_ denotes the regression coefficient for the edge from the node *x_k-1_* to *x_k_* in the CPD of *x_k_*, and *n* is the number of directed edges in the path ending at the outcome.

### Predictive Model from Bayesian Network

From the estimated Bayesian network, we derived a predictive model for %GSC. In a Bayesian network, a predictive model for an outcome variable given all the other variables is given as the conditional probability of the outcome variable given only the variables in the Markov blanket of the outcome, which consists of the parents, children, and co-parents of the children. This is because of the property of a Bayesian network that a variable is conditionally independent of all other variables given its Markov blanket. Since in our Bayesian network the Markov blanket of %GSC is composed solely of the parents of %GSC, we used the conditional probability of %GSC given its parents as the predictive model.

### Linear Regression Model

For comparison, a traditional multivariate linear regression model was estimated using the same dataset. The same 75 predictors that were used to estimate the Bayesian network were included in this model. The linear regression model was set up as

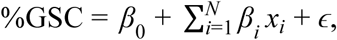

where *x_i_* represents the 𝑖th predictor (𝑖 = 1, . . ., 𝑁 for 𝑁 = 75), *β_0_* represents the intercept, *β_i_* denotes the regression coefficient for the 𝑖th predictor, and 𝜖 represents the residual error.

### Comparison of Predictive Models

To evaluate the prediction accuracy of the Bayesian network against the linear regression model, we performed 10-fold cross-validation. In each fold of the cross-validation, 90% of the samples were used as a training set to estimate each model and the remaining 10% were used as a test set to compute the mean squared error, defined as the average squared difference between the observed and predicted outcomes across the samples. The mean squared error averaged over 10 folds was used to assess the prediction accuracy of each model.

All analyses were conducted using the bnlearn package^47^ in R and the pgmpy package^48^ in Python.

## Results

Summary statistics of the samples and a comparison of those included and excluded have been previously reported.^12^ Among the 159 participants, the mean age was 74.98 years (SD = 4.51). The majority of the participants (62.9%) were women, most (91.2%) identified as White, most (88.7%) had attained more than a high school education, and just over half (52.8%) reported never smoking. On average, participants experienced -6.32% (SD = 5.22) slowing in gait speed from even to uneven surfaces.

### Bayesian Network Structure

Out of 75 predictors, 23 had Pearson correlation coefficients with %GSC greater or equal to 0.1 (**Table 1**). The hill-climbing algorithm applied to these 23 predictors and %GSC yielded a Bayesian network with 17 nodes that were connected to %GSC either directly or through a sequence of edges (**Figure 1**) and 6 nodes that were not connected to %GSC. The network structure and CPDs of the 17 connected nodes were retained in further analyses, including variables pertaining musculoskeletal and peripheral nervous systems, demographic, health, psychological well-being, and neuroimaging measures (**Supplemental Figure 1**). The 6 nodes excluded from further analysis were: right index finger vibration sensation (from PNS exam), wearing glasses, limbic striatal functional connectivity with anterior Brodmann area (BA) 24 in the right hemisphere, resting pulse rate, cognitive status, and physical activity.

**Table 1.**
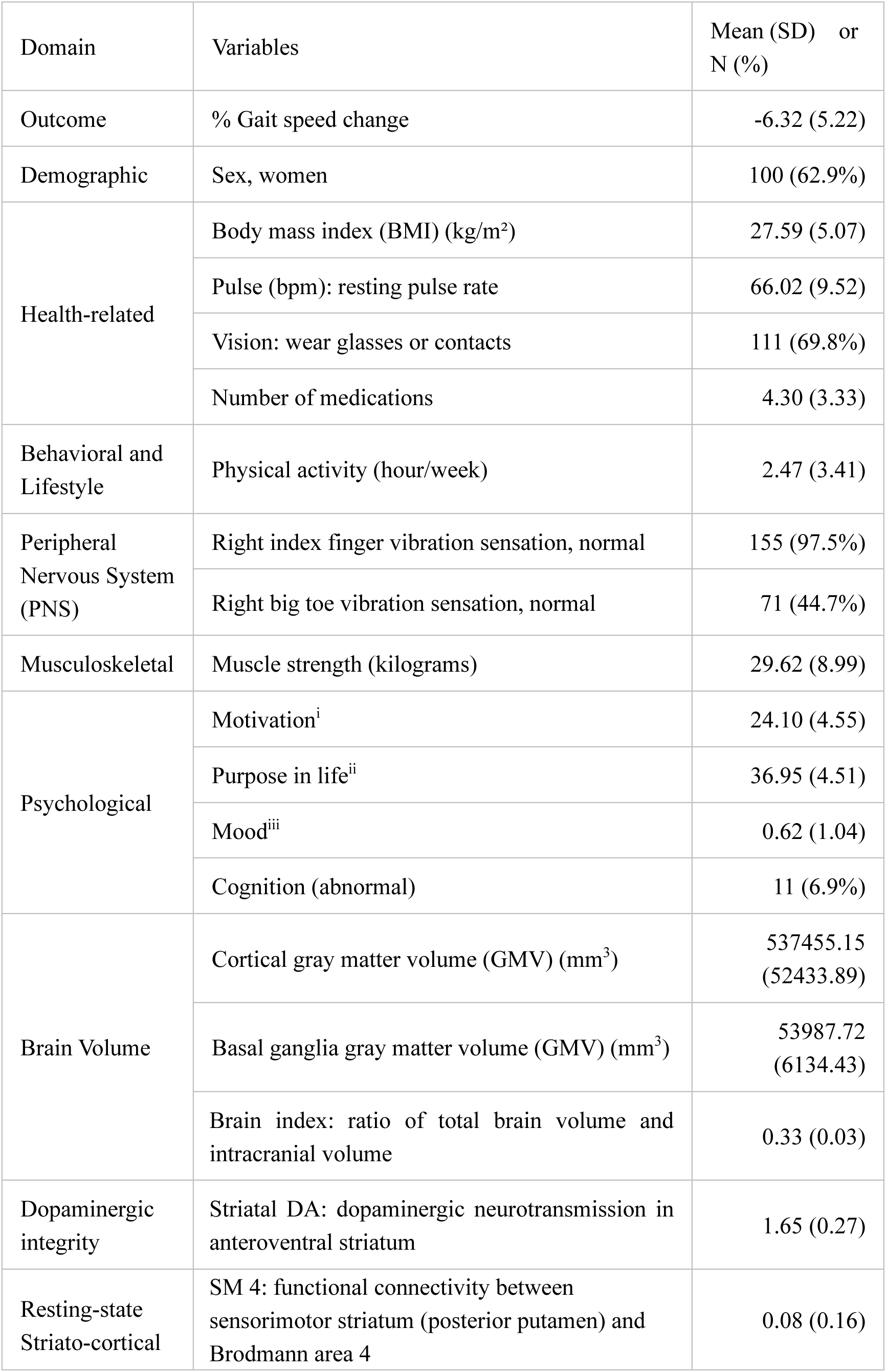
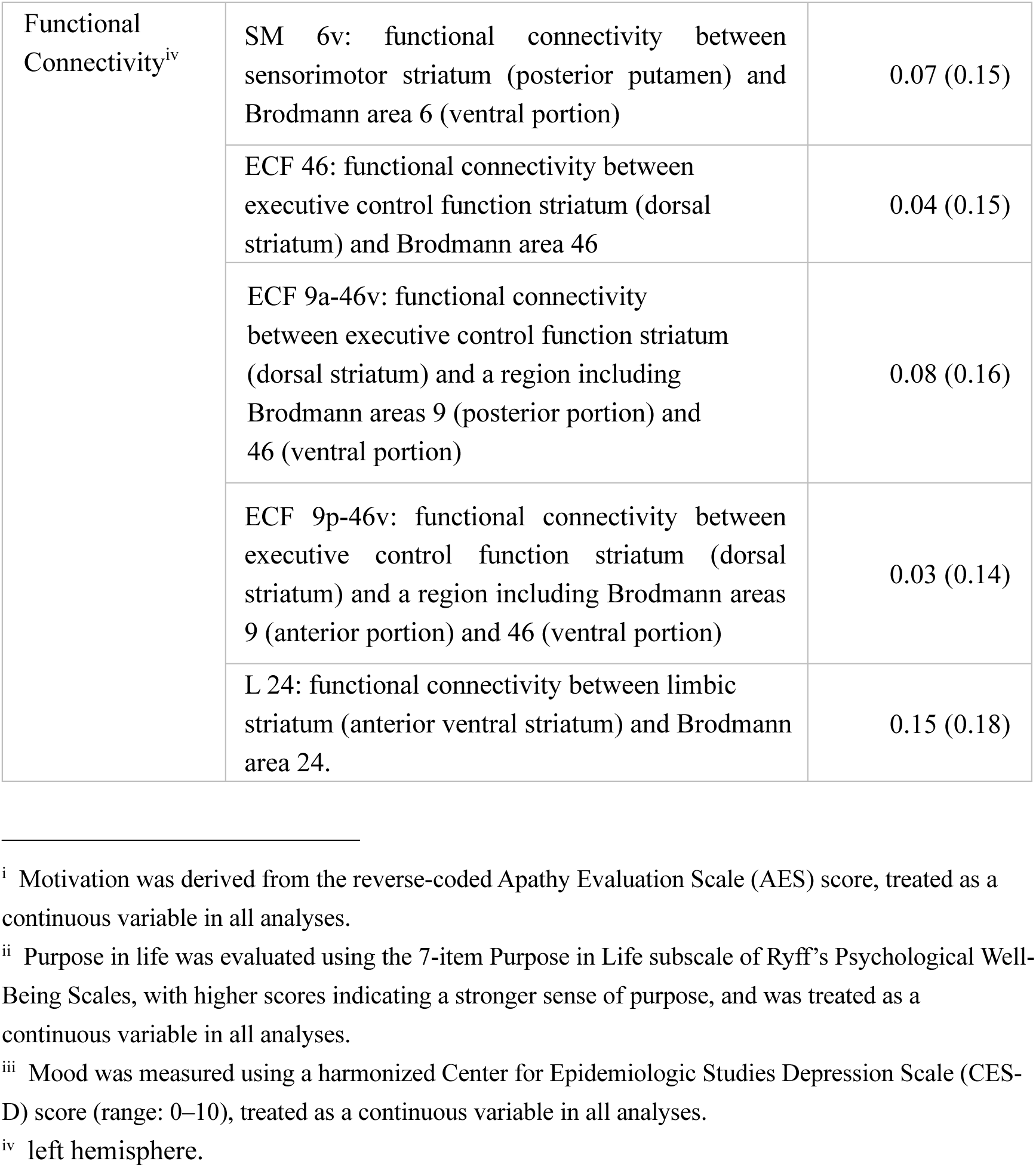
Sample characteristics for the 23 predictors and outcome included in the hill-climbing algorithm for Bayesian network estimation in N=159 participants.

**Figure 1.**
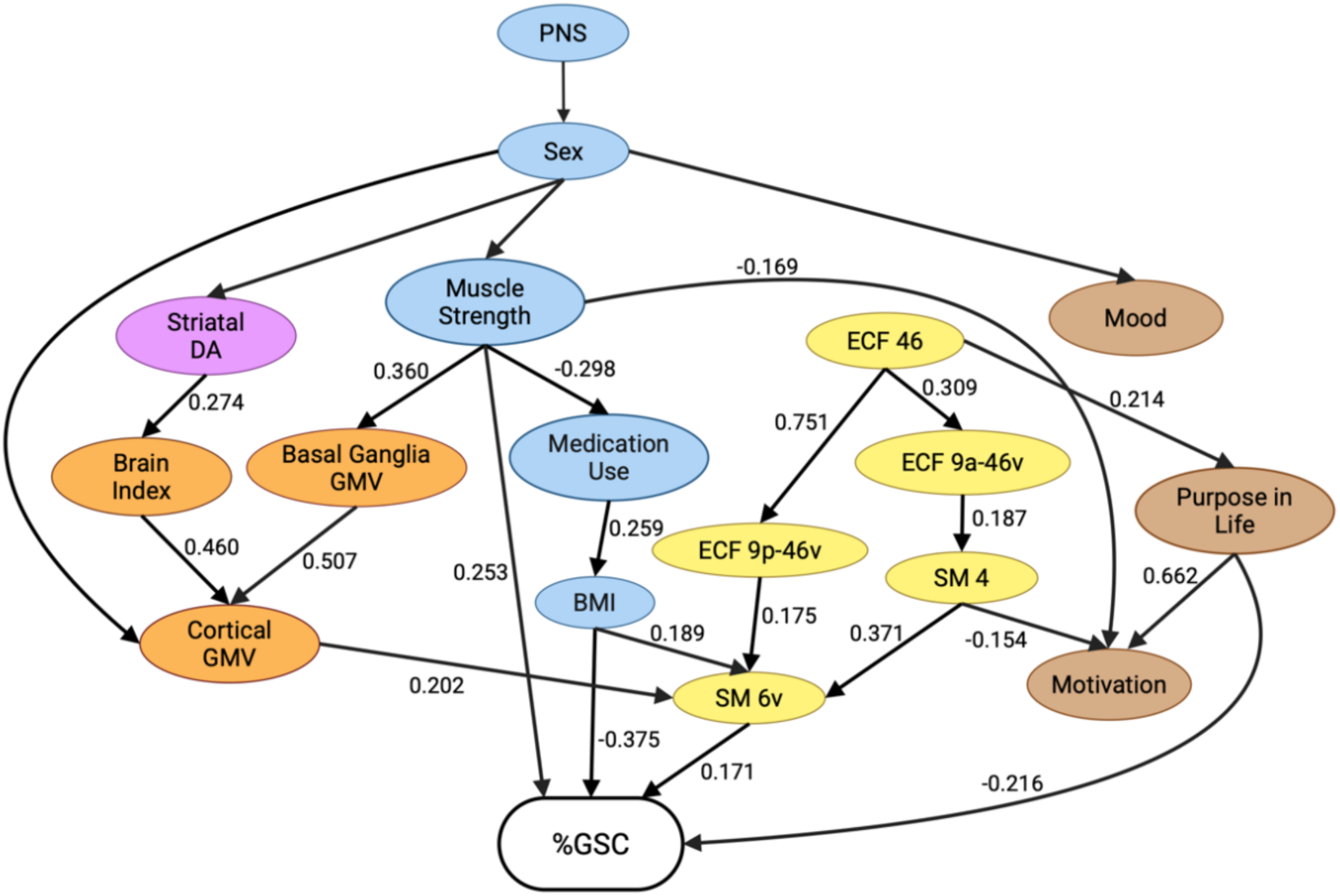
Bayesian network for % gait speed change (%GSC) when transitioning from even to uneven terrain. The nodes and edges of the estimated Bayesian network are shown for 17 nodes connected to %GSC through a sequence of edges. These 17 nodes included non-CNS features (blue nodes) as well as CNS features related to brain connectivity (yellow nodes), gray matter volume (orange nodes), psychosocial measures (brown nodes), and dopaminergic neurotransmission in the limbic striatum (purple node). PNS indicates the right index finger vibration sensation for proprioception. For continuous variables, the regression coefficients of their CPDs are shown with the corresponding edges. For continuous variables with sex as one of their parents, the CPD parameters for men are shown. See Supplemental Table 3 for the CPD parameters for discrete variables and for women.

The estimated network revealed an organization of the 17 nodes into clusters corresponding to distinct physiological systems of relevance for gait slowing. Specifically, neuroimaging measures were connected to each other in two clusters, one with functional connectivity measures (**Figure 1**, yellow) and the other with striatal DA/gray matter volume (**Figure 1**, purple and orange, respectively). The functional connectivity cluster included striato–cortical sensorimotor (SM) and executive control function (ECF) domains (ECF 46, ECF 9p-46v, ECF 9a-46v, SM 4, and SM 6v, left hemisphere). The other neuroimaging cluster included gray matter volumetric measures (brain index, and cortical and basal ganglia GMVs) and striatal dopaminergic neurotransmission in the limbic subregion. Psychological well-being features represented another cluster (purpose in life, motivation, and mood; **Figure 1**, brown). Other physiological systems in the network included muscle strength, BMI, medication use, proprioception from the PNS exam, and sex (**Figure 1**, blue).

### Model Comparison

The predictive performance of the Bayesian network was compared with that of a multivariate linear regression model using 10-fold cross-validation (**Figure 2**). Across all folds, the Bayesian network consistently demonstrated lower mean squared error than the linear regression. The mean squared error of the Bayesian network was 0.78 on average across folds, compared to 2.31 for the linear regression model. The lower mean squared errors with smaller variability across folds suggest that the Bayesian network provided more accurate and stable predictions of %GSC compared to the linear regression.

**Figure 2.**
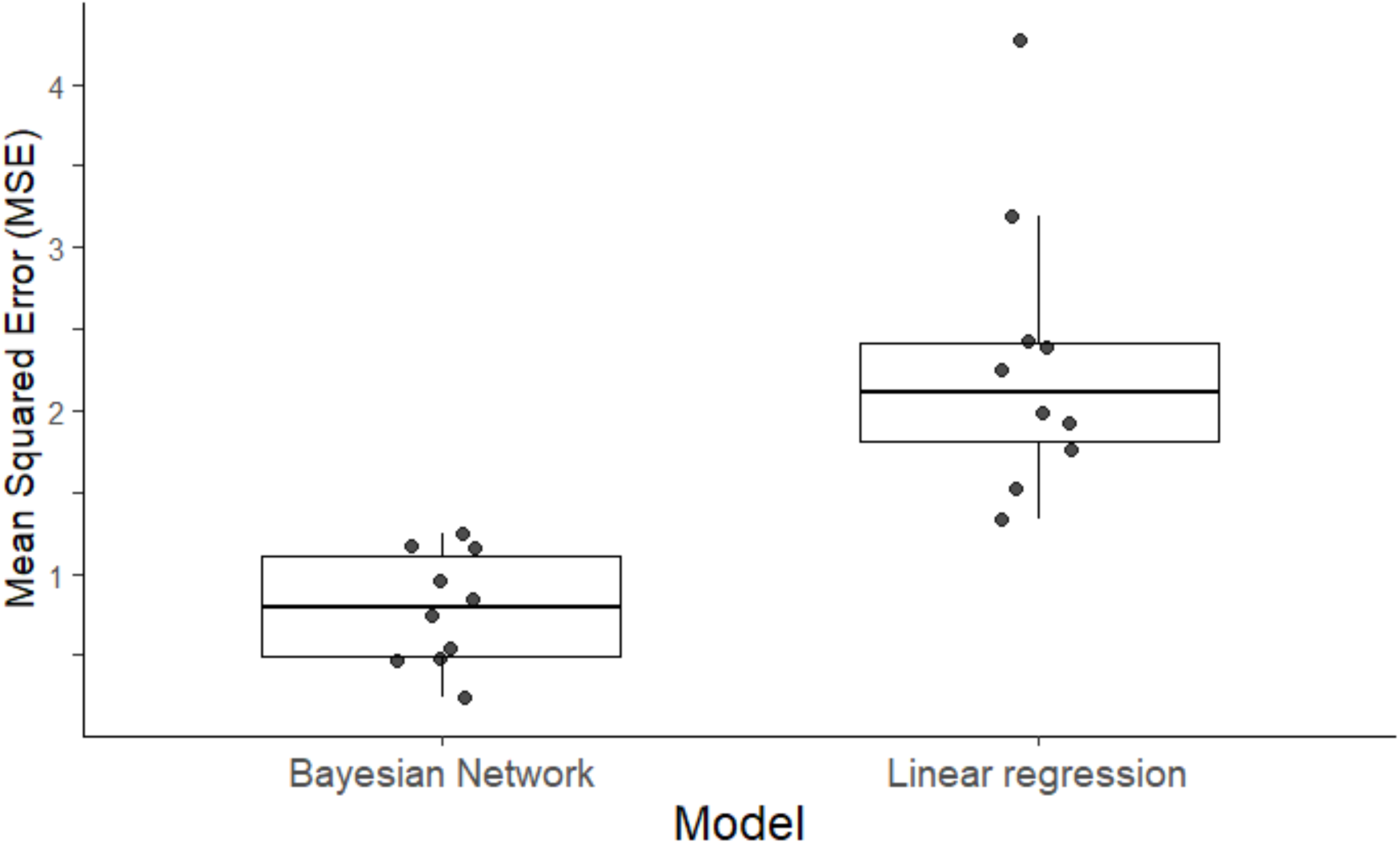
Comparison of predictive accuracy of the Bayesian network and multivariate linear regression. Boxplots show the distribution of mean squared errors (MSEs) across 10 folds of the cross-validation for each predictive model. Data points represent MSEs from individual folds. The MSE was obtained as the mean squared difference between the observed and predicted %GSCs across samples of the test set in each fold of the cross-validation. The Bayesian network has lower MSEs with smaller variability across folds than the linear regression model, indicating better predictive performance.

### Direct Paths Linking the Four Parent Nodes to %GSC

Predictors with direct paths to %GSC were evaluated using the parameters in the CPD of %GSC (**Figure 1**). There were four parents of %GSC with direct paths: BMI, muscle strength, SM 6v (functional connectivity between the sensorimotor striatum and the ventral portion of supplemental motor area BA 6), and purpose in life.

Among the four parent nodes, BMI had the direct effect of the largest absolute magnitude (*β*_BMI→%GSC_= -0.375). The negative sign of the association indicates that for higher BMI, there is less gait adaptability. Muscle strength had the highest positive effect among the nodes with direct effects (*β*_Muscle strength→%GSC_ = 0.253), indicating that for higher muscle strength there is better gait adaptability. SM 6v positively predicted %GSC (*β*_SM 6v→%GSC_= 0.171); this indicates that for higher SM striato-cortical functional connectivity there is better gait adaptability. Purpose in life had a negative direct effect on %GSC (*β*_Purpose in life→%GSC_= -0.216). The overall effect of the parent nodes was similar between men and women when stratified by sex (**Supplemental Table 2**).

### Indirect Paths Linking the Non-parent Nodes to % GSC via Parent Nodes

BMI and muscle strength influenced %GSC via indirect paths through other nodes, in addition to the direct paths. The contributions to %GSC through these indirect paths were evaluated based on the CPDs (**Figures 1** and **3**).

**Figure 3.**
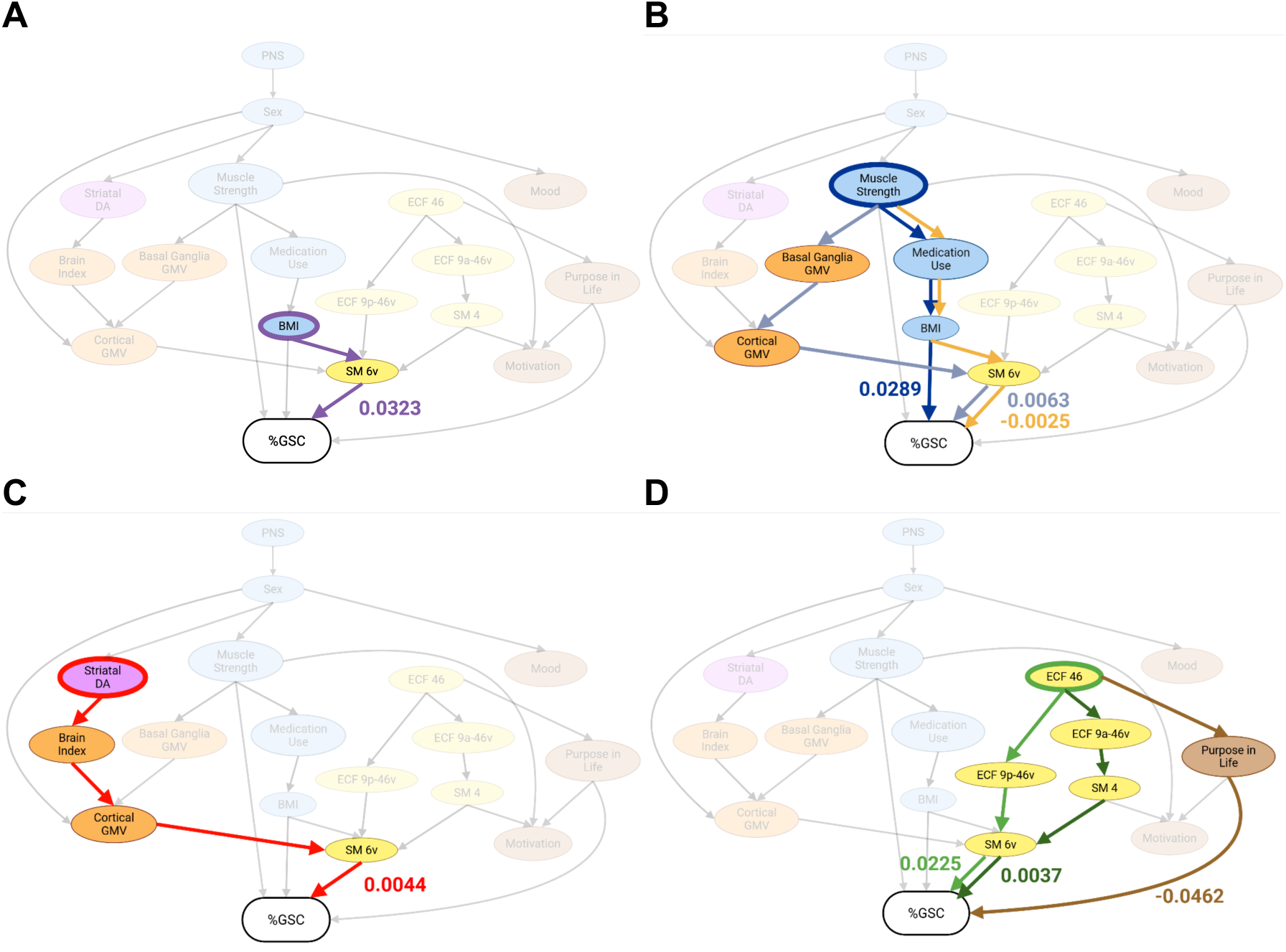
Indirect paths to %GSC in the Bayesian network. The indirect paths to %GSC and their path scores are shown for (**A**) BMI, (**B**) muscle strength, (**C**) striatal DA, and (**D**) ECF 46 as starting points. The sex-specific path scores for the paths through Cortical GMV in (**B**) and (**C**) were nearly identical between men and women.

BMI influenced %GSC indirectly via SM 6v (*β*_BMI→SM 6v_= 0.189) with positive path contribution (𝛾_BMI→ SM 6v→ %GSC_= 0.0323, **Figure 3A**). This positive indirect effect indicates that higher striato-cortical SM functional connectivity on this path diminished the negative direct impact of BMI on gait adaptability.

Muscle strength had three indirect paths to %GSC (**Figure 3B**). One path was through GMV of the basal ganglia (*β*_Muscle strength→Basal ganglia GMV_= 0.360) and from here to cortex and striato-cortical sensorimotor connectivity, resulting in positive contribution (𝛾Muscle strength→Basal ganglia GMV→Cortical GMV→SM 6v→%GSC = 0.0063, similar for women and men; **Figure 3B**, light blue). The other two indirect paths overlapped initially through medication use (*β*_Muscle strength→Medication use_= -0.298) and BMI (*β*_Medication use→BMI_= 0.259) with positive contribution of muscle strength to BMI (𝛾_Muscle strength→Medication use→BMI_ = -0.077), indicating higher muscle strength is associated with lower BMI. However, the two paths diverged from BMI: one path reached %GSC directly with positive contribution (𝛾_Muscle strength→Medication use→BMI→%GSC_ = 0.0289; **Figure 3B**, dark blue), indicating higher muscle strength positively impacts %GSC by lowering BMI, while the other path reached %GSC indirectly through striato-cortical SM 6v connectivity with negative contribution (𝛾_Muscle strength→Medication use→BMI→SM 6v→%GSC_= -0.0025; **Figure 3B**, yellow), indicating that the small protective effect of striato-cortical SM functional connectivity from the negative impact of BMI on %GSC (**Figure 3A**) is reduced as BMI is lowered by muscle strength. For all three paths, the indirect effects of muscle strength on %GSC were smaller compared to its direct effect. These indirect paths show that muscle strength influences %GSC not only via CNS factors (GMVs of basal ganglia and total brain) but also via non-CNS factor (burden of medical conditions and BMI).

### Other Indirect Paths to %GSC

Proprioception and sex appeared upstream in the network. Indirect paths from sex to %GSC were through three of the four child nodes of sex: striatal DA, cortical GMV, and muscle strength. The effect of sex on these features, as can be seen in the CPDs of these features (**Supplemental Table 3**), was consistent with sex-related differences in these measures. Men had greater muscle strength and cortical GMV but lower striatal DA than women; however, sex did not have significant influence on how these predictors influence other downstream variables including %GSC.

Striatal DA in the limbic subregion influenced %GSC indirectly (**Figure 3C**) through a cascade of positive associations through brain index (*β*_Striatal DA→Brain index_= 0.274), cortical GMV (*β*_Brain index→Cortical GMV_= 0.431 for women and 0.460 for men), and striato-cortical sensorimotor connectivity (*β*_Cortical GMV→SM 6v_= 0.202), for a total positive contribution of this path to %GSC (𝛾_Striatal DA→Brain index→Cortical GMV→SM 6v→%GSC_= 0.0041 for women and 0.0044 for men).

Striato-cortical functional connectivity with BA 46 (ECF 46) influenced %GSC through indirect paths engaging functional connectivity in ECF and SM striato-cortical networks, as well as purpose in life (**Figure 3D**). The functional connectivity path engaged two adjacent areas of ECF: the posterior portion of BA 9 and ventral BA 46 (ECF 9p-46v) (𝛾_ECF 46→ECF 9p-46v→SM 6v→%GSC_ = 0.0225; **Figure 3D**, light green); and the anterior portion of BA 9 and ventral BA 46 (ECF 9a-46v), which in turn influenced the striato-cortical SM connectivity with BA 4 (SM 4) (𝛾_ECF 46→ECF 9a-46v→SM 4→SM 6v→%GSC_ = 0.0037; **Figure 3D**, dark green). The path from ECF 46 through purpose in life showed that with greater functional connectivity of executive control regions, there are higher levels of perceived purpose in life (*β*_ECF 46→Purpose in life_ = 0.214), but negative path contribution on %GSC (𝛾_ECF 46→Purpose in life→%GSC_ = -0.0462; **Figure 3D**, brown).

## Discussion

In this study of older adults, the mean %GSC was negative, indicating that most individuals slowed when walking on the uneven compared with even surface, consistent with prior observations that uneven terrain challenges gait performance in aging populations.^5,12,49^ By applying a Bayesian network framework, we identified interrelationships between CNS and non-CNS features influencing %GSC either directly or indirectly through connections in the network. The predictive model for %GSC derived from the Bayesian network outperformed multivariate linear regression models in predictive accuracy, as shown by lower mean squared errors across the multiple folds in cross validation. Our results suggest the Bayesian network may serve as a platform for identifying interrelated key features dictating gait adaptability in aging populations; these features may be tested as targets of multi-modal interventions aimed at ameliorating age-related impairments in gait adaptability.

The predictive model constructed from the Bayesian network made predictions based solely on the four parents of %GSC in our network, indicating these features in the Markov blanket of the outcome carry all information necessary for prediction. We found direct connections between %GSC and variables spanning both CNS and non-CNS physiological domains: biomechanical factors (BMI), musculoskeletal function (muscle strength), psychological well-being (purpose in life), and neuroimaging integrity (sensorimotor functional connectivity). BMI and muscle strength emerged as the strongest predictors with opposite influences on %GSC, underscoring the role of maintaining healthier body composition and stronger skeletal muscle system to promote gait adaptability. Sensorimotor functional connectivity pointed to a neural substrate for adaptive walking, supporting the importance of this brain network for executing complex motor tasks. Higher purpose in life predicted a more negative %GSC, potentially reflecting the decision to walk more slowly when confronted with a challenging walking surface, perhaps to minimize risk of falls. This result seemingly contradicts other reports where higher purpose in life was related to faster gait.^50,51^ However, in these prior studies, gait speed was measured when walking at usual pace without added challenges or distractions, and associations with dynamic changes in gait speed during dual tasks have not been reported. The fact that these four predictors represent distinct but interconnected systems emphasizes that late-life gait adaptability is not determined by a single factor /domain but by the coordinated functioning of physical, psychological, and neural domains.

By modeling dependencies among features and separating direct from indirect effects through its graph structure, the Bayesian network mapped how upstream factors influenced outcomes via mediators. The network included several indirect paths of CNS and non-CNS features converging on %GSC, underscoring the importance of integrated peripheral and central features to orchestrate optimal motor control, and to maintain walking speed while negotiating a challenging surface.

Muscle strength influenced %GSC through indirect paths engaging neuroimaging and clinical features, in addition to its direct effect. Muscle strength was related to both higher GMV, and lower medication intake (a proxy of comorbidities) and BMI, before converging on sensorimotor connectivity to influence performance. This path linked together peripheral strength to central structure and health markers. On the other hand, BMI appeared to exert opposing influences: a strong negative direct association with performance, and a modest indirect improvement via greater sensorimotor functional connectivity. This result suggests higher sensorimotor functional connectivity in the presence of factors impacting locomotion, such as higher BMI, can represent an adaptive compensatory response, aimed at sustaining gait speed.

Sensorimotor connectivity is also of interest because it had several direct parents, thus appearing as a key node where brain structure, body composition, and ECF functional connectivity converged into a single signal directly related to performance. The network structure suggested a top-down control from executive networks feeding this sensorimotor hub, with an executive → sensorimotor cascade funneling into SM 6v. BA 6 including the premotor cortex and the supplemental motor area play key roles in planning, coordinating and sequencing complex movements. This may position executive control or motor planning as an upstream feature influencing sensorimotor coupling to negotiate uneven terrains. This dynamic sensorimotor integration appears to be “the last stop” before performance, working alongside ECF, biomechanical, and skeletomuscular control. Although the original dataset included striato–cortical connectivity measures from both left and right hemispheres, only left hemisphere connectivity variables exhibited sufficient correlation with %GSC to be included in the Bayesian network estimation with the hill-climbing algorithm, based on the pre-specified threshold (|r| > 0.1); one right hemisphere measure passed the threshold, but it remained unconnected in the network without any edges. This pattern may reflect the dominant role of the left hemisphere in voluntary movement, control, motor planning, and executive function, particularly in predominantly right-handed populations. Left hemisphere structures have been linked to greater vulnerability to age-related changes affecting mobility, particularly for tasks that require integration of executive and motor functions.^52^

Although striatal DA did not have a direct path to %GSC, there was an indirect path via other neural structures, indicating a neuromodulatory influence on gait speed from limbic dopaminergic neurotransmission via higher brain volumetric indices (higher brain index and greater cortical GMV) and sensorimotor connectivity. Notably, the striatal DA in this study was derived exclusively from the anterior ventral striatum, while DA neurotransmission from other striatal subregions did not appear connected to the gait-speed change or other factors influencing the gait-speed change directly or indirectly in the Bayesian network. This regional specificity suggests that reward-related dopaminergic signaling may be particularly relevant for gait adaptability in older adults, potentially reflecting vigor-related modulation of locomotion rather than primary motor execution. The total effect of this path on %GSC was comparable to the indirect effect of muscle strength through striato-cortical sensorimotor connectivity. Future studies should assess whether restoration of dopaminergic neurotransmission may promote gait adaptability. We and others have shown administration of small doses of DA-enhancing drugs such as L-DOPA is a feasible and safe strategy to improve gait speed in older adults with slow gait in the absence of neurological diseases.^53–55^ The beneficial effects of restoring DA neurotransmission may be due to improvement in functional connectivity. Modulating striatal DA influences functional connectivity in sensorimotor networks in PD,^56–60^ and healthy subjects,^61,62^ albeit whether these effects translate to improved adaptability has not been assessed.

These findings may inform potential future interventions. For example, multi-modal interventions to promote gait adaptability may test the effects of increasing functional connectivity of the sensorimotor network alongside strengthening exercises and weight controls. Strategies such as neuromuscular electrical stimulation (NMES) already offer such pleiotropic beneficial effects. Work done by us and others shows that NMES can elicit improvements in muscle structure and function similar to volitional exercise, with evidence that it improves walking performance in older adults.^63,64^ NMES elicits sensorimotor functional connectivity^65–72^ as well as neuroprotective effects and promotes brain perfusion and cognitive function.^72,73^

Negative results should also be noted. Although cognitive status was linked with physical activity, this relationship did not extend to %GSC, and neither appeared in the connected paths leading to %GSC, despite extensive literature linking both domains to mobility.^74,75^ Several factors may explain this pattern. First, the cognitive status measure in this study was a binary classification, and only 13 participants (6.2%) met criteria for possible mild cognitive impairment based on MoCA and CDR assessments. This limited variability likely reduced the ability to detect meaningful associations with %GSC. Second, once Bayesian network accounted for more proximal and stronger determinants, such as the four direct parents, the variance that cognitive status or physical activity might explain in traditional models may instead be captured by other neural or musculoskeletal pathways. Future studies with larger samples with greater ranges of cognitive function and physical activity are needed. Of note, results were similar in sex-stratified analyses. Sex appeared as an upstream variable in the estimated Bayesian network, directly influencing muscle strength, striatal DA, cortical GMV, and mood. This is consistent with well-established sex differences in musculoskeletal function, dopaminergic function, brain structure, and psychological well-being across aging.^76,77^ Although the CPDs of the child nodes of sex were stratified by sex and sex appeared to influence these child nodes in the estimated Bayesian network, there was little sex difference in how these child nodes of sex influence other downstream variables including %GSC. This finding suggests that sex primarily shapes the distribution of upstream factors and once these factors are accounted for, does not have strong direct effects on gait change.

Several limitations warrant consideration. First, the cross-sectional nature of the analysis limits causal inference; prospective studies are needed to confirm temporal ordering of identified pathways. Second, although the Bayesian network approach accommodates complex interdependencies, the model was built from a set of pre-selected variables, and unmeasured factors may have influenced results. Third, the modest sample size may have limited power to detect smaller effects through direct or indirect paths. However, our study has a relatively large sample for multimodal imaging research in aging and a very extensive neuroimaging phenotyping combining dopaminergic PET, structural MRI, and resting-state functional connectivity. The results of executive-to-sensorimotor connectivity pathway within the Bayesian network further support the robustness of our analytic methods.

In conclusion, our Bayesian network provides a framework to estimate the contribution of cross-system interactions to gait speed changes in response to challenging walking conditions. Our findings highlight modifiable targets for mobility interventions in older adults. The network-level influence of neurobiological variables such as striatal dopamine suggests that multi-domain interventions, such as combining physical training, cognitive engagement, and brain health promotion, may be most effective. Psychosocial engagement may also be a key pathway for preserving adaptability under challenging walking conditions. Future research should incorporate longitudinal designs, larger and more diverse samples, and interventional trials to test to what extent modifying combinations of upstream factors in these pathways may improve gait adaptability.

## Data Availability

All data produced in the present study are only available under license to approved researchers. Public access to the data is restricted because the data contains potentially sensitive participant information and consent limitations.

