## Supplemental Table 1 for "A Bayesian Network Analysis of Gait Speed Change Upon Transition to Uneven Surfaces in Older Adults"

**Supplemental Table 1. List of the 75 predictors included in the analysis.**

| Domain | Variable name | Description |
| --- | --- | --- |
| Demographic | Age | Age (years) |
|  | Sex | Sex (women, men) |
|  | Education | Years of education |
| Musculoskeletal System | Muscle strength | Maximal grip strength |
|  | Assistive walking device | Use of a walker or quad cane when they walk, self-report. |
|  | Joint pain | Presence of joint pain |
| Peripheral Nervous System Exam | Pronator drift motor exam (right side) | Assessment of upper-extremity motor control by observing for downward drift or pronation of the outstretched right arm (normal/abnormal) |
|  | Pronator drift motor exam (left side) | Assessment of upper-extremity motor control by observing for downward drift or pronation of the outstretched left arm (normal/abnormal) |
|  | Touch sensory exam (right side) | Assessment of light-touch sensation over the right lateral ankle (external malleolus) using a piece of cotton (normal/abnormal) |
|  | Touch sensory exam (left side) | Assessment of light-touch sensation over the left lateral ankle (external malleolus) using a piece of cotton (normal/abnormal) |
|  | Pin/Prick sensory exam (right side) | Assessment of pain sensation over the right lateral ankle (external malleolus) using a safety pin or sterile needle (normal/abnormal) |
|  | Pin/Prick sensory exam (left side) | Assessment of pain sensation over the left lateral ankle (external malleolus) using a safety pin or sterile needle (normal/abnormal) |
|  | Vibration sensory exam for index finger (right side) | Assessment of vibration sense at the distal interphalangeal joint of the right index finger using a tuning fork (normal/abnormal) |
|  | Vibration sensory exam for index finger (left side) | Assessment of vibration sense at the distal interphalangeal joint of the left index finger using a tuning fork (normal/abnormal) |
|  | Vibration sensory exam for big toe (right side) | Assessment of vibration sense at the distal interphalangeal joint of the right big toe using a tuning fork (normal/abnormal) |
|  | Vibration sensory exam for big toe (left side) | Assessment of vibration sense at the distal interphalangeal joint of the left big toe using a tuning fork (normal/abnormal) |
| Motor System | MDS UPDRS III | Unified Parkinson’s Disease Rating Scale – motor section |
|  | Finger-to-nose sensory exam (right side) | Assessment of upper-extremity coordination by having the participant alternately touch the examiner’s finger and their own nose with the right hand (normal/abnormal) |
|  | Finger-to-nose sensory exam (left side) | Assessment of upper-extremity coordination by having the participant alternately touch the examiner’s finger and their own nose with the left hand (normal/abnormal) |
|  | Finger-to-nose with eyes closed sensory exam (right side) | Assessment of right-arm coordination and proprioception while alternately touching the examiner’s finger and participant’s nose with eyes closed (normal/abnormal) |
|  | Finger-to-nose with eyes closed sensory exam (left side) | Assessment of left-arm coordination and proprioception while alternately touching the examiner’s finger and participant’s nose with eyes closed (normal/abnormal) |
|  | Romberg sensory exam (eyes open) | Assessment of balance by standing with feet together and eyes open for up to 30 seconds (normal/abnormal) |
|  | Romberg sensory exam (eyes closed) | Assessment of balance by standing with feet together and eyes closed for up to 30 seconds (normal/abnormal) |
| Psychological Well-being | Motivation | Derived from Apathy Evaluation Scale (total score) |
|  | Purpose in life | Ryff’s Purpose in Life subscale score |
|  | Energy level | Self-reported energy level in the past month |
|  | Physical fatigability | Pittsburgh Fatigability Scale for perceived physical fatigability |
|  | Mental fatigability | Pittsburgh Fatigability Scale for perceived mental fatigability |
|  | Mood | Harmonized depression variable from original and modified Center for Epidemiologic Studies Depression Scale (CES-D) |
| Cognition | Cognitive status | Classification of global cognitive function as normal cognition, mild cognitive impairment (MCI), or dementia, harmonized across cohorts using the Clinical Dementia Rating (CDR) scale and the Montreal Cognitive Assessment (MoCA). |
| Behavioral Lifestyle | Smoke status | Current smoking status (never, past, current) |
|  | Drinks per year | Self-reported estimated number of alcoholic drinks consumed per year |
|  | Physical activity | Weekly walking hours |
| Health | BMI | Body-mass index (kg/m²) |
|  | Systolic blood pressure | Mean systolic blood pressure |
|  | Diastolic blood pressure | Mean diastolic blood pressure |
|  | Resting pulse rate | Mean pulse rate |
|  | Medication use | Number of prescribed medications |
|  | Vision | Self-reported by participants on whether they usually wear glasses or contacts |
|  | Multimorbidity index | Self-reported medical history of cancer, atrial fibrillation, chronic kidney disease or renal failure, COPD, heart attack, heart failure, diabetes, stroke, and peripheral vascular disease |
| Resting State Functional Connectivity | Left ECF - Right ECF | Functional connectivity between the left and right dorsal striatum, important for executive control function |
|  | ECF 46, left | Functional connectivity between left dorsal striatum and Brodmann areas 46 in the left frontal cortex |
|  | ECF 9-46d, left | Functional connectivity between the left dorsal striatum and the Brodmann areas 9 and dorsal portions of Brodmann area 46 in the left frontal cortex |
|  | ECF 9a-46v, left | Functional connectivity between the left dorsal striatum and the anterior portions of Brodmann area 9 and the ventral portion of Brodmann area 46 in the left frontal cortex |
|  | ECF 9p-46v, left | Functional connectivity between the left dorsal striatum and the posterior portion of Brodmann area 9 and the ventral portion of Brodmann area 46 in the left frontal cortex |
|  | ECF 46, right | Functional connectivity between the right dorsal striatum and Brodmann area 46 in the right frontal cortex |
|  | ECF 9-46d, right | Functional connectivity between the right dorsal striatum and the Brodmann areas 9 and dorsal portions of Brodmann area 46 in the right frontal cortex |
|  | ECF 9a-46v, right | Functional connectivity between the right dorsal striatum and the anterior portions of Brodmann area 9 and the ventral portion of Brodmann area 46 in the right frontal cortex |
|  | ECF 9p-46v, right | Functional connectivity between the right dorsal striatum and the posterior portion of Brodmann area 9 and the ventral portion of Brodmann area 46 in the right frontal cortex |
|  | Left SM - Right SM | Functional connectivity between left and right sensorimotor striatum (posterior putamen) |
|  | SM 4, left | Functional connectivity between left posterior putamen and Brodmann areas 4 in the left frontal cortex |
|  | SM 6v, left | Functional connectivity between left posterior putamen and the ventral portion of Brodmann areas 6 in the left frontal cortex |
|  | SM 4, right | Functional connectivity between right posterior putamen and Brodmann areas 4 in the right frontal cortex |
|  | SM 6v, right | Functional connectivity between right posterior putamen and the ventral portion of Brodmann areas 6 in the right frontal cortex |
|  | Left limbic - Right limbic | Functional connectivity between left and right limbic striatum (anterior ventral striatum). |
|  | Limbic 24a, left | Functional connectivity between left anterior ventral striatum and the anterior portion of Brodmann areas 24 in the left frontal cortex |
|  | Limbic OFC, left | Functional connectivity between left anterior ventral striatum and the left orbitofrontal cortex |
|  | Limbic OFCp, left | Functional connectivity between left anterior ventral striatum and the left posterior orbitofrontal cortex |
|  | Limbic 32s, left | Functional connectivity between left anterior ventral striatum and the superior portion of Brodmann area 32 in the left frontal cortex |
|  | Limbic 24a, right | Functional connectivity between right anterior ventral striatum and the anterior portion of Brodmann areas 24 in the right frontal cortex |
|  | Limbic OFC, right | Functional connectivity between right anterior ventral striatum and the right orbitofrontal cortex |
|  | Limbic OFCp, right | Functional connectivity between right anterior ventral striatum and the right posterior orbitofrontal cortex |
|  | Limbic 32s, right | Functional connectivity between right anterior ventral striatum and the superior portion of Brodmann area 32 in the right frontal cortex |
| Brain Structure (MRI) | Total WMH | Total white matter hyperintensities in mm^3^ |
|  | Normalized WMH | Normalized total white matter hyperintensities in mm^3^ |
|  | Brain index | Ratio of total brain volume and intracranial volume |
|  | Basal ganglia GMV | Basal ganglia gray-matter volume |
|  | Cortical GMV | Total cortical gray-matter volume |
| Striatal dopaminergic neurotransmission (PET) | Anterior ventral striatum | [^11^C]DTBZ binding potential in anterior ventral striatum |
|  | Pre dorsal caudate | [^11^C]DTBZ binding potential in pre dorsal caudate |
|  | Post dorsal caudate | [^11^C]DTBZ binding potential in post dorsal caudate |
|  | Anterior putamen | [^11^C]DTBZ binding potential in anterior putamen |
|  | Posterior putamen | [^11^C]DTBZ binding potential in posterior putamen |
|  | Associative striatum | [^11^C]DTBZ binding potential in anterior putamen, pre dorsal caudate, post dorsal caudate |
|  | Striatum | [^11^C]DTBZ binding potential in striatum |
