## Supplemental Figure 1 for "A Bayesian Network Analysis of Gait Speed Change Upon Transition to Uneven Surfaces in Older Adults"

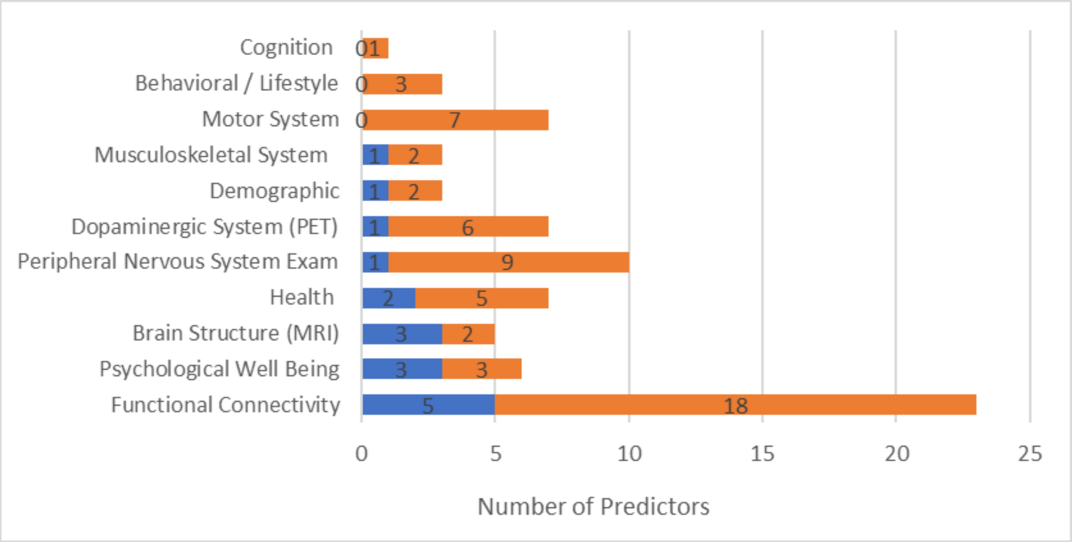


**Supplemental Figure 1**. **Predictors with or without paths to %GSC in the estimated Bayesian network.** Among the 75 harmonized variables (excluding the outcome) included in the analysis, those that were connected to %GSC through paths with one or more edges in the Bayesian network (blue) are compared with those that were not connected to %GSC (orange).
