## Supplemental Table 2 for "A Bayesian Network Analysis of Gait Speed Change Upon Transition to Uneven Surfaces in Older Adults"

**Supplemental Table 2. Overall effects of parents of %GSC on %GSC in the Bayesian Network stratified by sex.**

| Predictor | Men | Women |
| --- | --- | --- |
| BMI | −0.3586 | −0.3586 |
| Muscle strength | 0.2860 | 0.2860 |
| SM 6v | 0.1088 | 0.1084 |
| Purpose in life | −0.2102 | −0.2102 |
