## Supplemental Table 3 for "A Bayesian Network Analysis of Gait Speed Change Upon Transition to Uneven Surfaces in Older Adults"

**Supplemental Table 3. The CPD parameter estimates in the Bayesian network.**

| Variable | CPD parameters |
| --- | --- |
| ECF 46 | $\text{β}_{\text{ECF 46,0}}$ = 0.023  $\text{σ}_{\text{ECF 46}}$ = 0.997 |
| ECF 9a–46v | $\text{β}_{\text{ECF 9a-46v,0}}$ = 0.011  $\text{β}_{\text{ECF 46→ECF 9a-46v}}$ = 0.309  $\text{σ}_{\text{ECF 9a-46v}}$ = 0.961 |
| ECF 9p–46v | $\text{β}_{\text{ECF 9p-46v,0}}$ = 0.009  $\text{β}_{\text{ECF 46→ECF 9p-46v}}$ = 0.751  $\text{σ}_{\text{ECF 9p-46v}}$ = 0.656 |
| SM 4 | $\text{β}_{\text{SM 4,0}}$ = 0.056  $\text{β}_{\text{ECF 9a-46v→SM 4}}$ = 0.187  $\text{σ}_{\text{SM 4}}$ = 0.987 |
| SM 6v | $\text{β}_{\text{SM 6v,0}}$ = 0.110  $\text{β}_{\text{SM 4→SM 6v}}$ = 0.371  $\text{β}_{\text{ECF 9p-46v→SM 6v}}$ = 0.175  $\text{β}_{\text{BMI→SM 6v}}$ = 0.189  $\text{β}_{\text{Cortical GMV→SM 6v}}$ = 0.202  $\text{σ}_{\text{SM 6v}}$ = 0.885 |
| Basal ganglia GMV | $\text{β}_{\text{Basal ganglia GMV,0}}$ = 0.008  $\text{β}_{\text{Muscle }\text{strength→Basal}\text{ ganglia GMV}}$ = 0.360  $\text{σ}_{\text{Basal ganglia GMV}}$ = 0.979 |
| Brain index | $\text{β}_{\text{Brain index,0}}$ = −0.059  $\text{β}_{\text{Striatal }\text{DA→Brain}\text{ index}}$ = 0.274  $\text{σ}_{\text{Brain index}}$ = 0.961 |
| Medication use | $\text{β}_{\text{Medication use,0}}$ = −0.010  $\text{β}_{\text{Muscle }\text{strength→Medication}\text{ use}}$ = −0.298  $\text{σ}_{\text{Medication use}}$ = 0.973 |
| BMI | $\text{β}_{\text{BMI,0}}$ = −0.136  $\text{β}_{\text{Medication }\text{use→BMI}}$ = 0.259  $\text{σ}_{\text{BMI}}$ = 0.896 |
| Purpose in life | $\text{β}_{\text{Purpose in life,0}}$ = 0.009  $\text{β}_{\text{ECF 46→Purpose in life}}$ = 0.214  $\text{σ}_{\text{Purpose in life}}$ = 0.949 |
| %GSC | $\text{β}_{\text{\%GSC,0}}$ = −0.038  $\text{β}_{\text{SM 6v→\%GSC}}$ = 0.171  $\text{β}_{\text{BMI→\%GSC}}$ = −0.375  $\text{β}_{\text{Purpose in life→\%GSC}}$ = −0.216  $\text{β}_{\text{Muscle strength→\%GSC}}$ = 0.253  $\text{σ}_{\text{\%GSC}}$ = 0.797 |
| Motivation | $\text{β}_{\text{Motivation}\text{,}\text{0}}$ = −0.050  $\text{β}_{\text{SM 4→Motivation}}$ = −0.154  $\text{β}_{\text{Putpose}\text{ in }\text{life→Motivation}}$ = 0.662  $\text{β}_{\text{Muscle }\text{strength→Motivation}}$ = 0.169  $\text{σ}_{\text{Motivation}}$ = 0.750 |
| Cortical GMV (Sex = Man) | $\text{β}_{\text{Cortical GMV}\text{,0}}$ = 0.563  $\text{β}_{\text{Basal ganglia }\text{GMV→Cortical}\text{ GMV}}$ = 0.507  $\text{β}_{\text{Brain }\text{index→Cortical}\text{ GMV}}$ = 0.460  $\text{σ}_{\text{Cortical GMV}}$ = 0.720 |
| Cortical GMV (Sex = Woman) | $\text{β}_{\text{Cortical GMV}\text{,0}}$ = −0.334  $\text{β}_{\text{Basal }\text{ganglig}\text{ }\text{GMV→Cortical}\text{ GMV}}$ = 0.512  $\text{β}_{\text{Brain }\text{index→Cortical}\text{ GMV}}$ = 0.431  $\text{σ}_{\text{Cortical GMV}}$ = 0.597 |
| Striatal DA (Sex = Man) | $\text{β}_{\text{Striatal DA,0}}$ = −0.277  $\text{σ}_{\text{Striatal DA}}$ = 0.948 |
| Striatal DA (Sex = Woman) | $\text{β}_{\text{Striatal DA,0}}$ = 0.250  $\text{σ}_{\text{Striatal DA}}$ = 1.009 |
| Mood (Sex = Man) | $\text{β}_{\text{Mood,0}}$ = −0.225  $\text{σ}_{\text{Mood}}$ = 0.671 |
| Mood (Sex = Woman) | $\text{β}_{\text{Mood,0}}$ = 0.109  $\text{σ}_{\text{Mood}}$ = 1.043 |
| Physical activity (Cognition = Normal) | $\text{β}_{\text{Physical activity}\text{,0}}$ = 0.039  $\text{σ}_{\text{Physical activity}}$ = 0.965 |
| Physical activity (Cognition = MCI or Dementia) | $\text{β}_{\text{Physical activity}\text{,0}}$ = 0.638  $\text{σ}_{\text{Physical activity}}$ = 1.997 |
| Muscle strength (Sex = Man) | $\text{β}_{\text{Muscle strength,0}}$ = 0.924  $\text{σ}_{\text{Muscle strength}}$ = 0.826 |
| Muscle strength (Sex = Woman) | $\text{β}_{\text{Muscle strength,0}}$ = −0.563  $\text{σ}_{\text{Muscle strength}}$ = 0.591 |
| Right index finger vibration | P(Normal) = 0.025  P(Abnormal) = 0.975 |
| Right big toe vibration | P(Normal) = 0.554  P(Abnormal) = 0.446 |
| Sex (Right big toe vibration = Normal) | P(Man\|Normal) = 0.489  P(Woman\|Normal) = 0.511 |
| Sex (Right big toe vibration = Abnormal) | P(Man\|Abnormal) = 0.225  P(Woman\|Abnormal) = 0.775 |
| Cognition | P(Mild cognitive impairment) = 0.931  P(Dementia) = 0.069 |
| Vision | P(No glass) = 0.302  P(Wear glasses) = 0.698 |

Notes:

β₍_Parent→Child_₎ denotes the conditional regression coefficient relating the parent node to the child node.
β₀ denotes the intercept term of the conditional Gaussian distribution for the child node.
σ denotes the residual standard deviation of the child node.

For discrete nodes, P(State) denotes the marginal probability of each state estimated from the Bayesian network.
